# Human leukocyte antigen susceptibility map for SARS-CoV-2

**DOI:** 10.1101/2020.03.22.20040600

**Authors:** Austin Nguyen, Julianne K. David, Sean K. Maden, Mary A. Wood, Benjamin R. Weeder, Abhinav Nellore, Reid F. Thompson

## Abstract

Genetic variability across the three major histocompatibility complex (MHC) class I genes (human leukocyte antigen [HLA] A, B, and C) may affect susceptibility to and severity of severe acute respiratory syndrome 2 (SARS-CoV-2), the virus responsible for coronavirus disease 2019 (COVID-19). We execute a comprehensive *in silico* analysis of viral peptide-MHC class I binding affinity across 145 HLA -A, -B, and -C genotypes for all SARS-CoV-2 peptides. We further explore the potential for cross-protective immunity conferred by prior exposure to four common human coronaviruses. The SARS-CoV-2 proteome is successfully sampled and presented by a diversity of HLA alleles. However, we found that HLA-B*46:01 had the fewest predicted binding peptides for SARS-CoV-2, suggesting individuals with this allele may be particularly vulnerable to COVID-19, as they were previously shown to be for SARS (1). Conversely, we found that HLA-B*15:03 showed the greatest capacity to present highly conserved SARS-CoV-2 peptides that are shared among common human coronaviruses, suggesting it could enable cross-protective T-cell based immunity. Finally, we report global distributions of HLA types with potential epidemiological ramifications in the setting of the current pandemic.

**IMPORTANCE:** Individual genetic variation may help to explain different immune responses to a virus across a population. In particular, understanding how variation in HLA may affect the course of COVID-19 could help identify individuals at higher risk from the disease. HLA typing can be fast and inexpensive. Pairing HLA typing with COVID-19 testing where feasible could improve assessment of viral severity in the population. Following the development of a vaccine against SARS-CoV-2, the virus that causes COVID-19, individuals with high-risk HLA types could be prioritized for vaccination.

## INTRODUCTION

Recently, a new strain of betacoronavirus (severe acute respiratory syndrome coronavirus 2, or SARS-CoV-2) has emerged as a global pathogen, prompting the World Health Organization in January 2020 to declare an international public health emergency (2). In the large coronavirus family, comprising enveloped positive-strand RNA viruses, SARS-CoV-2 is the seventh encountered strain that causes respiratory disease in humans (3) ranging from mild -- the common cold -- to severe -- the zoonotic Middle East Respiratory Syndrome (MERS-CoV) and Severe Acute Respiratory Syndrome (SARS-CoV). As of April 2020, there are over one million presumed or confirmed cases of coronavirus disease 19 (COVID-19) worldwide, with total deaths exceeding 50,000 (4). While age and many comorbidities, including cardiovascular and pulmonary disease, appear to increase the severity and mortality of COVID-19 (5–10), approximately 80% of infected individuals have mild symptoms (11). As with SARS-CoV (12, 13) and MERS-CoV (14, 15), children seem to have low susceptibility to the disease (16–18); despite similar infection rates as adults (19) only 5.9% of pediatric cases are severe or critical, possibly due to lower binding ability of the ACE2 receptor in children or generally higher levels of antiviral antibodies (20). Other similarities (21–23) including genomic (24, 25) and immune system response (26–34) between SARS-CoV-2 and other coronaviruses (35), especially SARS-CoV and MERS-CoV, are topics of ongoing active research, results of which may inform an understanding of the severity of infection (36) and improve the ongoing work of immune landscape profiling (37–41) and vaccine discovery (29, 38, 42–49).

Human leukocyte antigen (HLA) alleles, which are critical components of the viral antigen presentation pathway, have been shown in previous studies to confer differential viral susceptibility and severity of disease. For instance, disease caused by the closely-related SARS-CoV (24, 25) shows increased severity among individuals with the HLA-B*46:01 genotype (1). Associations between HLA genotype and disease severity extend broadly to several other unrelated viruses. For example, in human immunodeficiency virus 1 (HIV-1), certain HLA types (e.g. HLA-A*02:05) may reduce risk of seroconversion (50), and in dengue virus, certain HLA alleles (e.g. HLA-A*02:07, HLA-B*51) are associated with increased secondary disease severity among ethnic Thais (51).

While a detailed clinical picture of the COVID-19 pandemic continues to emerge, there remain substantial unanswered questions regarding the role of individual genetic variability in the immune response against SARS-CoV-2 (51). We hypothesize that individual HLA genotypes may differentially induce the T-cell mediated antiviral response, and could potentially alter the course of disease and its transmission. In this study, we performed a comprehensive *in silico* analysis of viral peptide-MHC class I binding affinity, across 145 different HLA types, for the entire SARS-CoV-2 proteome.

## RESULTS

To explore the potential for a given HLA allele to produce an antiviral response, we assessed the HLA binding affinity of all possible 8-to 12-mers from the SARS-CoV-2 proteome (n=48,395 unique peptides). We then removed from further consideration 16,138 peptides that were not predicted to enter the MHC class I antigen processing pathway via proteasomal cleavage. For the remaining 32,257 peptides, we repeated binding affinity predictions for a total of 145 different HLA types, and we show here the SARS-CoV-2-specific distribution of per-allele proteome presentation (predicted binding affinity threshold <500nM, Figure 1, Supplementary Table S1). Importantly, we note that the putative capacity for SARS-CoV-2 antigen presentation is unrelated to the HLA allelic frequency in the population (Figure 1). We identify HLA-B*46:01 as the HLA allele with the fewest predicted binding peptides for SARS-CoV-2. We performed the same analyses for the closely-related SARS-CoV proteome (Supplementary Figure S1) and similarly note that HLA-B*46:01 was predicted to present the fewest SARS-CoV peptides, keeping with previous clinical data associating this allele with severe disease (1).

**Figure 1:**
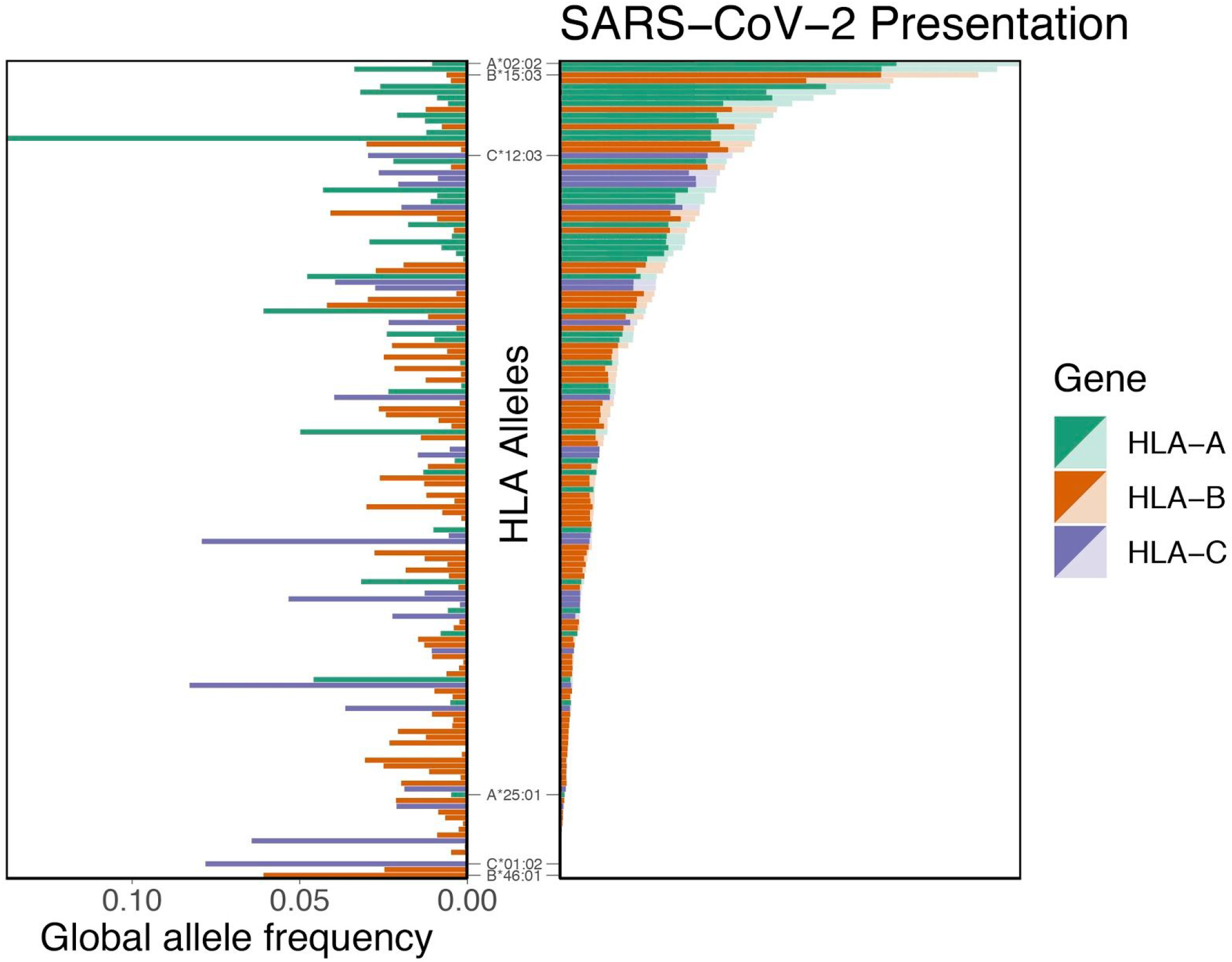
Distribution of HLA allelic presentation of 8-to 12-mers from the SARS-CoV-2 proteome. At right, the number of peptides (see Supplementary Table S1) that putatively bind to each of 145 HLA alleles is shown as a series of horizontal bars, with dark and light shading indicating the number of tightly (<50nM) and loosely (<500nM) binding peptides respectively, and with green, orange, and purple colors representing HLA-A, -B, and -C alleles, respectively. Alleles are sorted in descending order based on the number of peptides they bind (<500nM). The corresponding estimated allelic frequency in the global population is also shown (to left), with length of horizontal bar indicating absolute frequency in the population.

To assess the potential for cross-protective immunity conferred by prior exposure to common human coronaviruses (i.e. HKU1, OC43, NL63, and 229E), we next sought to characterize the conservation of the SARS-CoV-2 proteome across diverse coronavirus subgenera to identify highly conserved linear epitopes. After aligning reference proteome sequence data for 5 essential viral components (ORF1ab, S, E, M, and N proteins) across 34 distinct alpha- and betacoronaviruses, including all known human coronaviruses, we identified 48 highly conserved amino acid sequence spans (Appendix 1). Acknowledging the challenges inferring cross-protective immunity among closely related peptides, we confined attention exclusively to identical peptide matches. Among conserved sequences, 44 SARS-CoV-2 sequences would each be anticipated to generate at least one 8-to 12-mer linear peptide epitope also present within at least one other common human coronavirus (Supplementary Table S2, Figure 2). In total, 564 such 8-to 12-mer peptides were found to share 100% identity with corresponding OC43, HKU1, NL63, and 229E sequences (467, 460, 179, and 157 peptides, respectively) (Supplementary Table S3).

**Figure 2:**
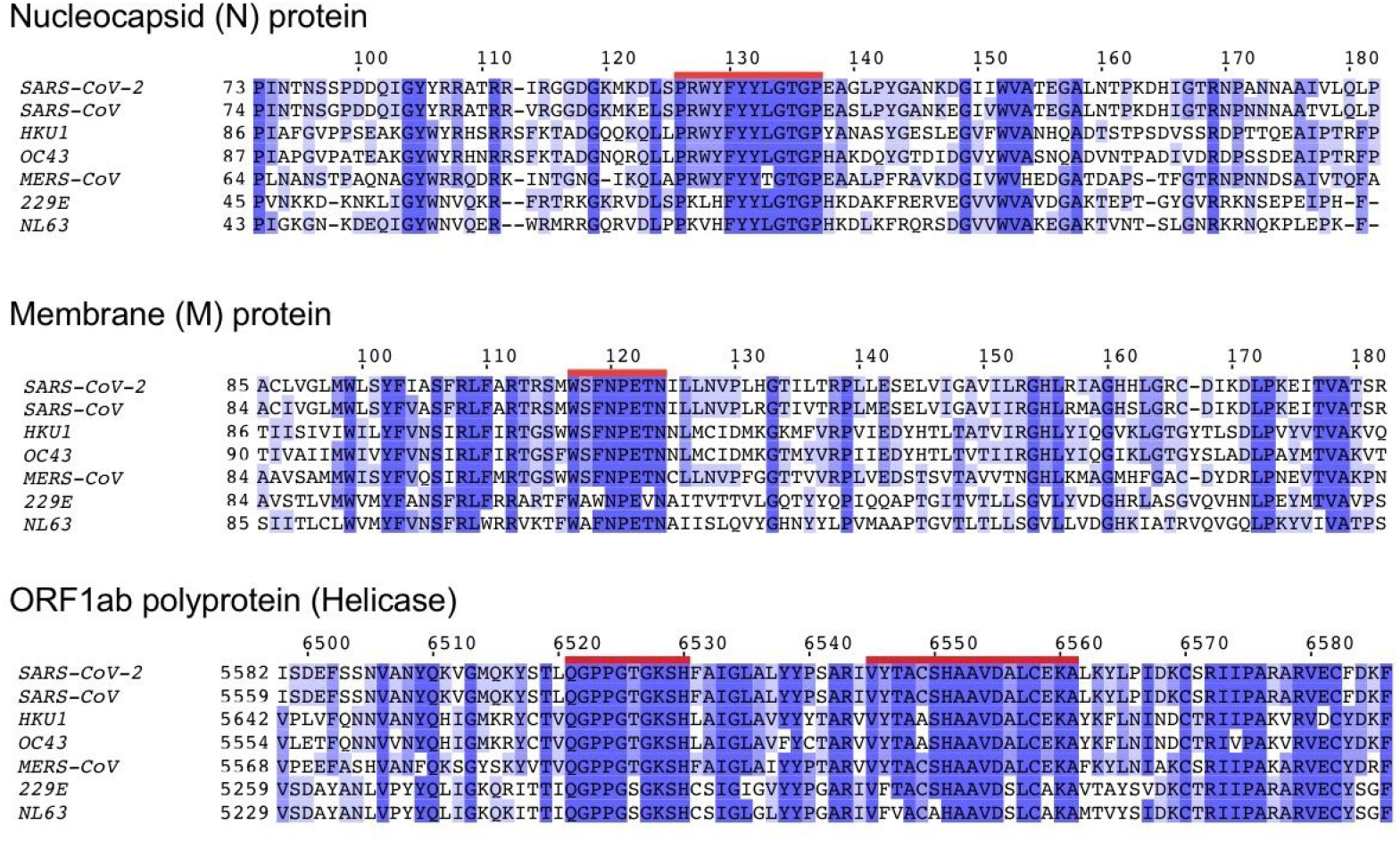
Amino acid sequence conservation of four linear peptide example sequences from three human coronavirus proteins. Protein sequence alignments are shown for nucleocapsid (N), membrane (M), and ORF1ab polyprotein (Helicase) across all five known human betacoronaviruses (SARS-CoV-2, SARS-CoV, HKU1, OC43, and MERS-CoV) and two known human alphacoronaviruses (229E and NL63). Each row in the three depicted sequence alignments corresponds to the protein sequence from the indicated coronavirus, with starting coordinate of the viral protein sequence shown at left and position coordinates of the overall alignment displayed above. Blue shading indicates the extent of sequence identity, with the darkest blue shading indicating 100% match for that amino acid across all sequences. The four red highlighted sequences correspond to highly conserved peptides ≥8 amino acids in length (PRWYFYYLGTGP, WSFNPETN, QPPGTGKSH, VYTACSHAAVDALCEKA, see Supplementary Table S2).

For the subset of these potentially cross-protective peptides that are anticipated to be generated via the MHC class I antigen processing pathway, we performed binding affinity predictions across 145 different HLA-A, -B, and -C alleles. As above, we demonstrate the SARS-CoV-2-specific distribution of per-allele presentation for these conserved peptides. We found that alleles HLA-A*02:02, HLA-B*15:03, and HLA-C*12:03 were the top presenters of conserved peptides. Conversely, we note that 56 different HLA alleles demonstrated no appreciable binding affinity (<500nM) to any of the conserved SARS-CoV-2 peptides, suggesting a concomitant lack of potential for cross-protective immunity from other human coronaviruses. We note, in particular, HLA-B*46:01 is among these alleles. Note also that the putative capacity for conserved peptide presentation is unrelated to the HLA allelic frequency in the population (Figure 3). Moreover, we see no appreciable global correlation between conservation of the SARS-CoV-2 proteome and its predicted MHC binding affinity, suggesting a lack of selective pressure for or against the capacity to present coronavirus epitopes (p=0.27 [Fisher’s exact test], Supplementary Figure S2).

**Figure 3:**
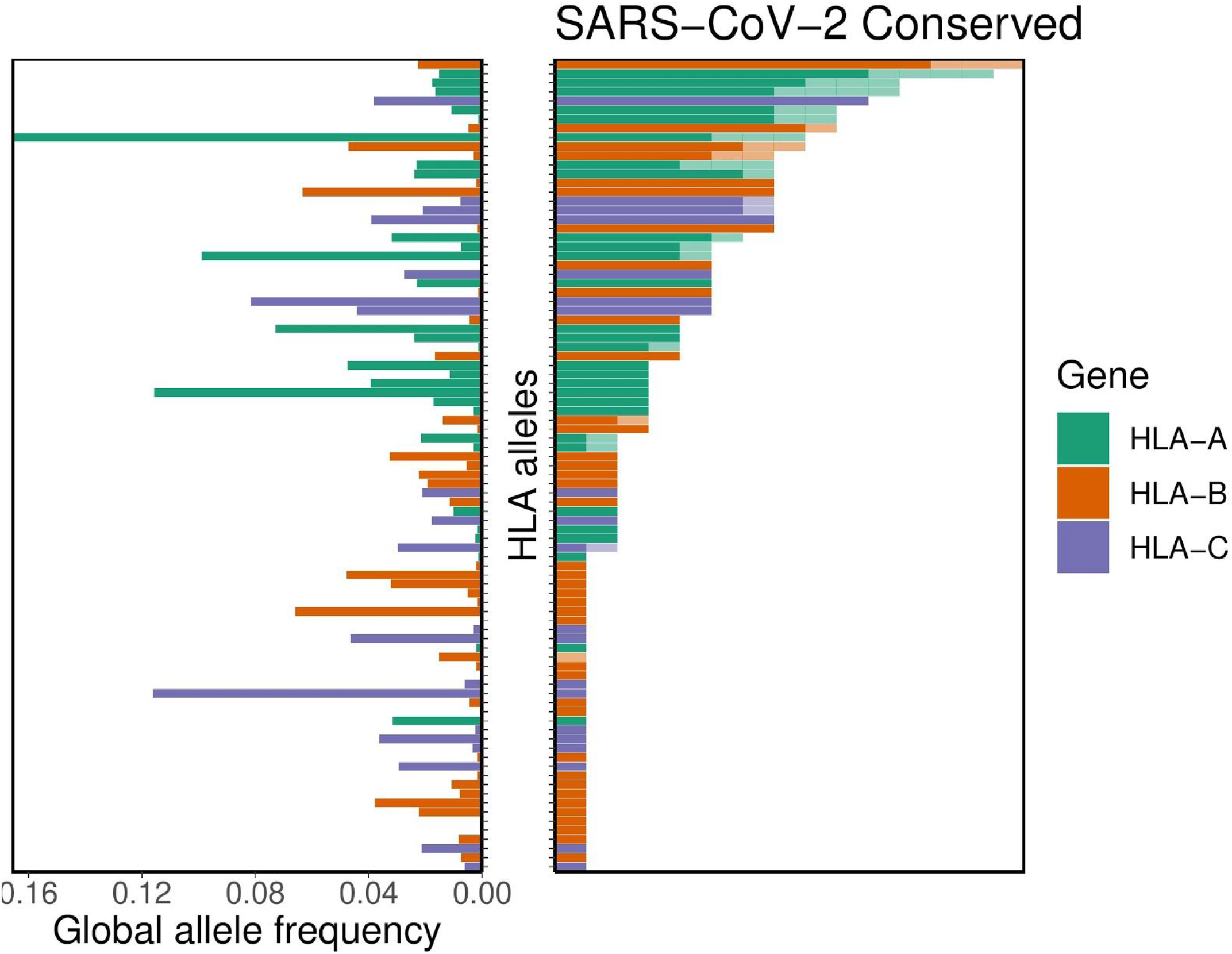
Distribution of HLA allelic presentation of highly conserved human coronavirus peptides with potential to elicit cross-protective immunity to COVID-19. At right, the number of conserved peptides (see Supplementary Table S3) that putatively bind to a subset of 89 HLA alleles is shown as a series of horizontal bars, with dark and light shading indicating the number of tightly (<50nM) and loosely (<500nM) binding peptides, respectively, and with green, orange, and purple colors representing HLA-A, -B, and -C alleles, respectively. Alleles are sorted in descending order based on the number of peptides they are anticipated to present (binding affinity <500nM). The corresponding allelic frequency in the global population is also shown (to left), with length of horizontal bar indicating absolute frequency in the population.

We were further interested in whether certain regions of the SARS-CoV-2 proteome showed differential presentation by the MHC class I pathway. Accordingly, we surveyed the distribution of antigen presentation capacity across the entire proteome, highlighting its most conserved peptide sequences (Figure 4). Throughout the entire proteome, HLA-A and HLA-C alleles exhibited the relative largest and smallest capacity to present SARS-CoV-2 antigens, respectively. However each of the three major class I genes exhibited a very similar pattern of peptide presentation across the proteome (Supplementary Figure S3). We additionally note that peptide presentation appears to be independent of estimated time of peptide production during viral life cycle, with indistinguishable levels of peptide presentation of both early and late SARS-CoV-2 peptides (Supplementary Figure S4).

**Figure 4:**
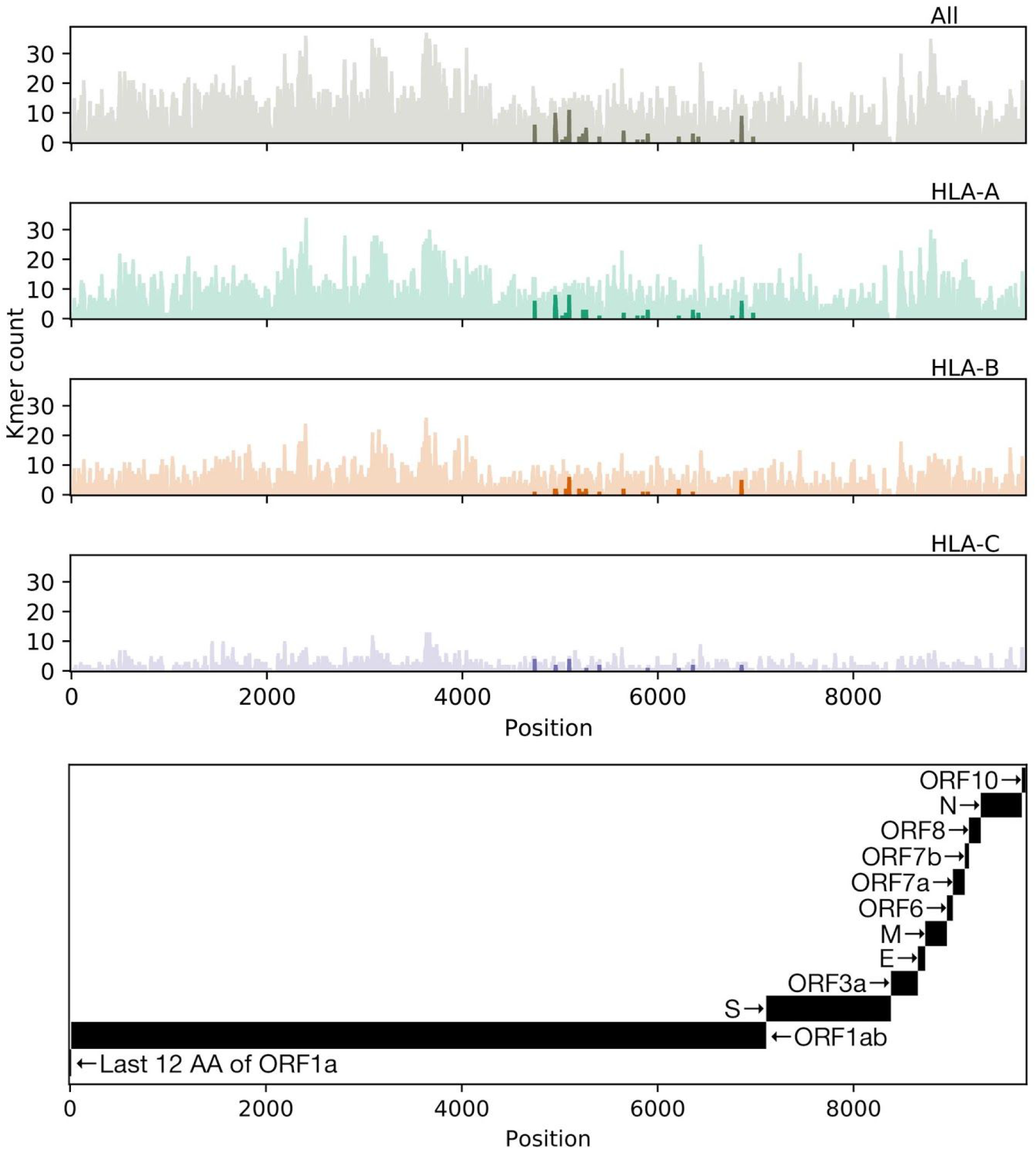
Distribution of allelic presentation of conserved 8-to 12-mers across the entire SARS-CoV-2 proteome for all HLA alleles and individually for HLA-A, HLA-B, and HLA-C (first, second, third, and fourth plots from top, respectively) with dark and light shading indicating the number of tightly (<50nM) and loosely (<500nM) binding peptides, respectively. Positions are derived from a concatenation of coding sequences (CDSes) indicated in the bottom panel. Tightly binding peptides are confined to ORF1ab. The sequence begins with only the last 12 amino acids of ORF1a because all but the last four amino acids of ORF1a are contained in ORF1ab, and we considered binding peptides up to 12 amino acids in length.

Given the global nature of the current COVID-19 pandemic, we sought to describe population-level distributions of the HLA alleles best (and least) able to generate a repertoire of SARS-CoV-2 epitopes in support of a T-cell based immune response. While we present global maps of individual HLA allele frequencies for the full set of 145 different alleles studied herein (Appendix 2), we specifically highlight the global distributions of the three best-presenting (A*02:02, B*15:03, C*12:03) and three of the worst-presenting (A*25:01, B*46:01, C*01:02) HLA-A, -B, and -C alleles (Figure 5). Note that all allelic frequencies are aggregated by country, but implicitly reflect the distribution of HLA data available on the Allele Frequency Net Database (52).

**Figure 5:**
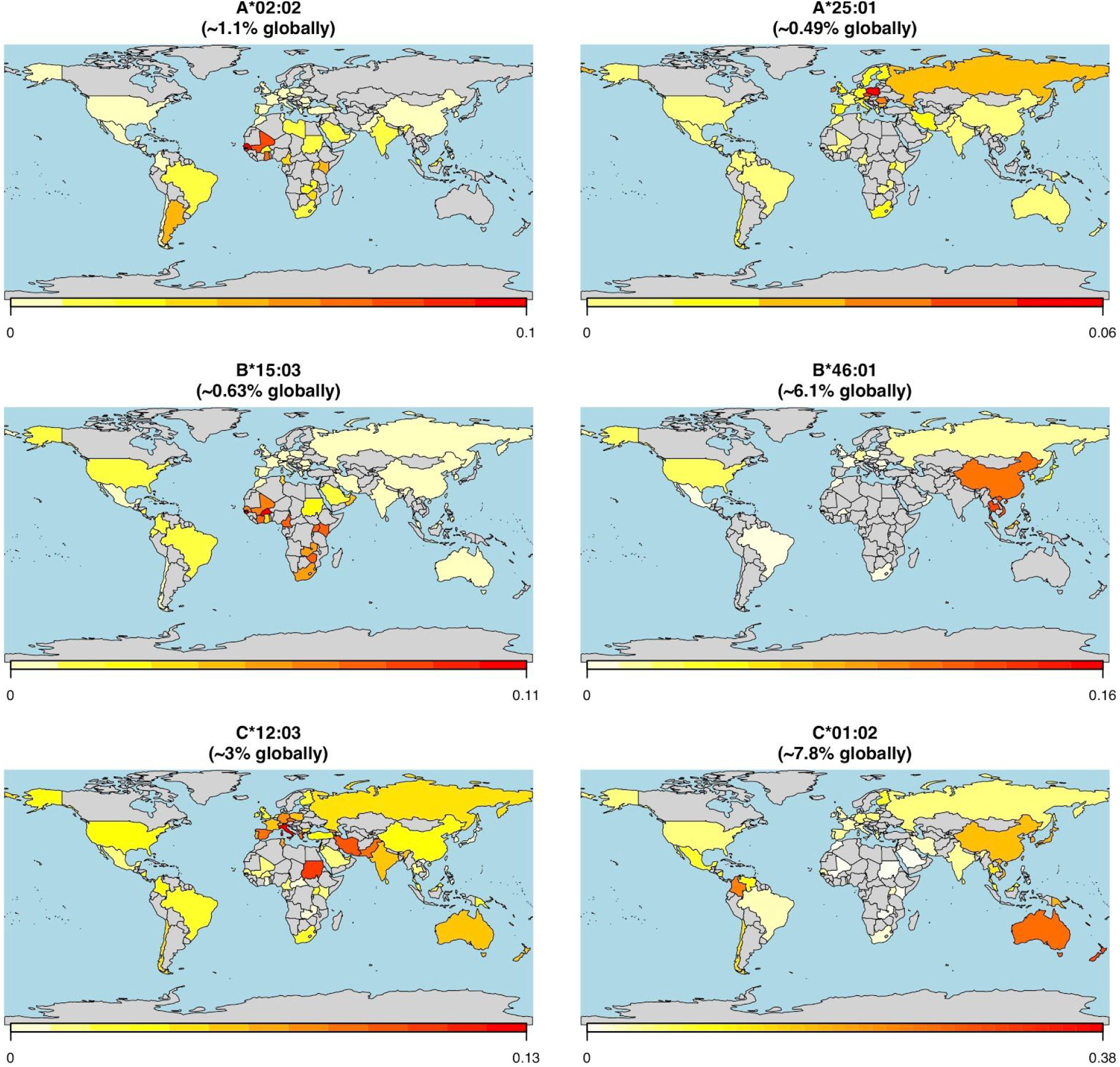
Global HLA allele frequency distribution heatmaps for six HLA-A, -B, and -C alleles. The leftmost panels show the global allele frequency distributions by country for three representative alleles (HLA-A*02:02, HLA-B*15:03, and HLA-C*12:03) with the predicted capacities to present the greatest repertoire of epitopes from the SARS-CoV-2 proteome (21.1%, 19.1%, and 7.9% of presentable epitopes, respectively). Conversely, the rightmost panels show the global allele frequency distributions by country for three representative alleles (HLA-A*25:01, HLA-B*46:01, and HLA-C*01:02) with the least predicted epitope presentation from the SARS-CoV-2 proteome (0.2%, 0%, 0% of presentable epitopes, respectively). Heatmap color corresponds to the individual HLA allele frequency within each country ranging from least (white/yellow) to most (red) frequent as indicated in the legend below each map.

Finally, we acknowledge that nearly all individuals have two HLA-A/B/C haplotypes constituting as few as three but as many as six distinct alleles, potentially buffering against the lack of presentation from a single poorly-presenting allele. We sought to describe whether allele-specific variability in SARS-CoV-2 presentation extends to full HLA haplotypes and to whole individual HLA genotypes. For six representative alleles with the highest (HLA-A*02:02, HLA-B*15:03, and HLA-C*12:03) and lowest (HLA-A*25:01, HLA-B*46:01, and HLA-C*01:02) predicted capacity for SARS-CoV-2 epitope presentation, these differences remain significant at the haplotype level, albeit with wide variability in presentation among different haplotypes (Figure 6).

**Figure 6:**
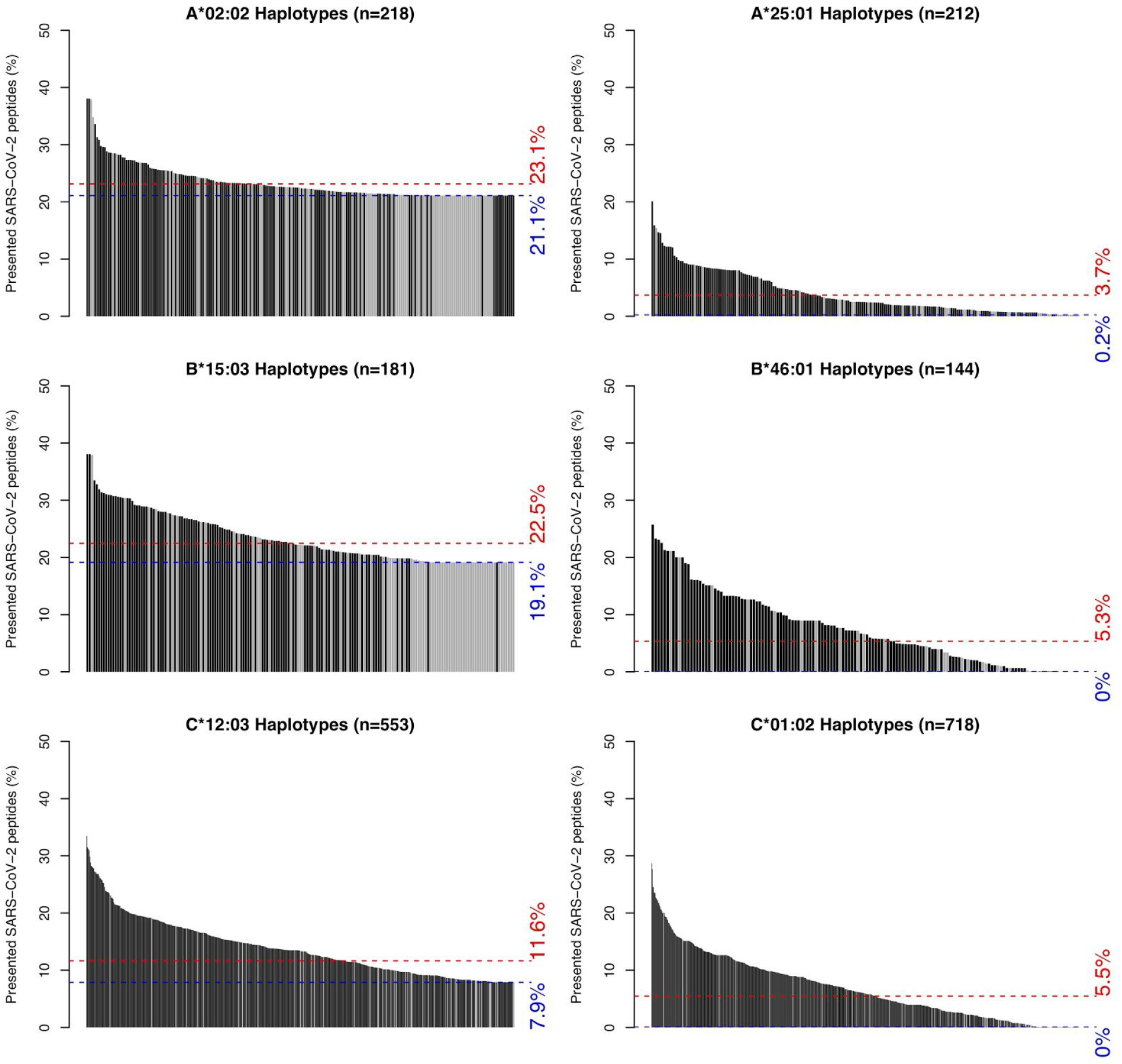
Distributions of SARS-CoV-2 peptide presentation across HLA haplotypes. The leftmost panels show the distributions of SARS-CoV-2 peptide presentation capacity for haplotypes containing one of three representative HLA alleles (HLA-A*02:02, HLA-B*15:03, and HLA-C*12:03) with the greatest predicted repertoire of epitopes from the SARS-CoV-2 proteome. Conversely, the rightmost panels show the distributions of SARS-CoV-2 peptide presentation capacity for haplotypes containing one of three representative alleles (HLA-A*25:01, HLA-B*46:01, and HLA-C*01:02) with the least predicted epitope presentation from the SARS-CoV-2 proteome. Black and gray bars represent full or partial haplotypes, respectively. Blue and red Dashed lines represent the percent of presented SARS-CoV-2 peptides for the indicated allele itself (blue) and its global population frequency weighted average presentation across its observed haplotypes (red).

Haplotype-level data for all 145 alleles is included in Supplementary Figure S5 and Appendix 2. We then identified 3,382 individuals with full HLA genotype data and noted wide variability in their capacity to present peptides from the SARS-CoV-2 proteome, albeit with a small minority of individuals at either extreme (Supplementary Figure S6).

## DISCUSSION

To the best of our knowledge, this is the first study to evaluate per-allele viral proteome presentation across a wide range of HLA alleles using MHC-peptide binding affinity predictors. This study also introduces the relationship between coronavirus sequence conservation and MHC class I antigen presentation. We show that individual HLA, haplotype, and full genotype variability likely influence the capacity to respond to SARS-CoV-2 infection, and we note certain alleles in particular (e.g. HLA-B*46:01) that could be associated with more severe infection, as previously shown with SARS-CoV (1). Indeed, we further compare SARS-CoV and

SARS-CoV-2 peptide presentation and note a high degree of similarity between the two across HLA types. Finally, this is the first study to report global distributions of HLA types and haplotypes with potential epidemiological ramifications in the setting of the current pandemic. We found that in general, there is no correlation between the HLA allelic frequency in the population and allelic capacity to bind SARS-CoV or SARS-CoV-2 peptides, irrespective of estimated timing of peptide production during the viral replication cycle. While we are not aware of any studies explicitly reporting the relationship between human coronavirus epitope abundance and immune response, there is data in vaccinia virus that suggests that early peptide antigens are more likely to generate CD8+ T-cell responses while antibody and CD4+ T-cell responses are more likely to target later mRNA expression with higher peptide abundance in the virion (53).

We note, however, several limitations to this our work. First and foremost, while we note that a handful of our binding affinity predictions were borne out in experimentally validated SARS-CoV peptides (Supplementary Table S4), we acknowledge that this is an entirely *in silico* study. As we are unable to obtain individual-level HLA typing and clinical outcomes data for any real-world COVID-19 populations at this time, the data presented is theoretical in nature, and subject to many of the same limitations implicit to the MHC binding affinity prediction tool(s) upon which it is based. As such, we are unable to assess the relative importance of HLA type compared to known disease-modifying risk factors such as age and clinical comorbidities (5–10). We further note that peptide-MHC binding affinity is limited as a predictor of subsequent T-cell responses (54–56), and we do not study T-cell responses herein. As such, we are ill-equipped to explore phenomena such as original antigenic sin (57–59), where prior exposure to closely related infection(s) may trigger T-cell anergy (60–62) or immunopathogenesis (63) in the setting of a novel infection. We explored only a limited set of 145 well-studied HLA alleles, but note that this analysis could be performed across a wider diversity of genotypes (49). Additionally, we did not assess genotypic heterogeneity or *in vivo* evolution of SARS-CoV-2, which could modify the repertoire of viral epitopes presented, or otherwise modulate virulence in an HLA-independent manner (64, 65) (https://nextstrain.org/ncov). We also do not address the potential for individual-level genetic variation in other proteins (e.g. angiotensin converting enzyme 2 [ACE2] or transmembrane serine protease 2 [TMPRSS2], essential host proteins for SARS-CoV-2 priming and cell entry (66) to modulate the host-pathogen interface.

Unless and until the findings we present here are clinically validated, they should not be employed for any clinical purposes. However, we do at this juncture recommend integrating HLA testing into clinical trials and pairing HLA typing with COVID-19 testing where feasible to more rapidly develop and deploy predictor(s) of viral severity in the population, and potentially to tailor future vaccination strategies to genotypically at-risk populations. This approach may have additional implications for the management of a broad array of other viruses.

## MATERIALS AND METHODS

### Sequence retrieval and alignments

Full polyprotein 1ab (ORF1ab), spike (S) protein, membrane (M) protein, envelope (E) protein, and nucleocapsid (N) protein sequences were obtained for each of 34 distinct but representative alpha and betacoronaviruses from a broad genus and subgenus distribution, including all known human coronaviruses (i.e. SARS-CoV, SARS-CoV-2, MERS-CoV, HKU1, OC43, NL63, and 229E). FASTA-formatted protein sequence data (full accession number list available in Supplementary Table S4S5) was retrieved from the National Center of Biotechnology Information (NCBI) (67). For each protein class (i.e. ORF1ab, S, M, E, N), all 34 coronavirus sequences were aligned using the Clustal Omega v1.2.4 multisequence aligner tool employing the following parameters: sequence type [Protein], output alignment format [clustal_num], dealign [false], mBed-like clustering guide-tree [true], mBed-like clustering iteration [true], number of combined iterations [0], maximum guide tree iterations [-1], and maximum HMM iterations [-1] (68). For the purposes of estimating time of viral peptide production, we classified ORF1a and ORF1b peptides as “early” while all other peptides produced by subgenomic mRNAs were classified as “late” (69, 70).

### Conserved peptide assessment

Aligned sequences were imported into Jalview v. 2.1.1 (71) with automated generation of the following alignment annotations: 1) sequence consensus, calculated as the percentage of the modal residue per column, 2) sequence conservation (0-11), measured as a numerical index reflecting conservation of amino acid physico-chemical properties in the alignment, 3) alignment quality (0-1), measured as a normalized sum of BLOSUM62 ratios for all residues at each position, 4) occupancy, calculated as the number of aligned residues (not including gaps) for each position. In all cases, sequence conservation was assessed for each of three groups: only human coronaviruses (n=7), all betacoronaviruses (n=16), and combined alpha- and betacoronavirus sequences (n=34). Aligned SARS-CoV-2 sequence and all annotations were manually exported for subsequent analysis. Conserved human coronavirus peptides were defined as those with a length ≥8 consecutive amino acids, each with an agreement of SARS-CoV-2 and ≥4 other human coronavirus sequences with the consensus sequence (Supplementary Table S2). For each of these conserved peptides, we also assessed the component number of 8-to 12-mers sharing identical amino acid sequence between SARS-CoV-2 and each of the four other common human coronaviruses (i.e. OC43, HKU1, NL63, 229E) (Supplementary Table S3). For all peptides, human, beta, and combined conservation scores were obtained using a custom R v.3.6.2 script as the mean sequence conservation (minus gap penalties where relevant) (see https://github.com/pdxgx/covid19).

### Peptide-MHC class I binding affinity predictions

FASTA-formatted input protein sequences from the entire SARS-CoV-2 and SARS-CoV proteomes were obtained from NCBI RefSeq database (67) under accession numbers NC_045512.2 and NC_004718.3. We kmerized each of these sequences into 8-to 12-mers to assess MHC class I-peptide binding affinity across the entire proteome. MHC class I binding affinity predictions were performed using 145 different HLA alleles for which global allele frequency data was available as described previously (72) (see Supplementary Table S1 S5) with netMHCpan v4.0 (73) using the ‘-BA’ option to include binding affinity predictions and the ‘-l’ option to specify peptides of lengths 8-12 (Supplementary Table S1). Binding affinity was not predicted for peptides containing the character ‘|’ in their sequences. Additional MHC class I binding affinity predictions were performed on all 66 MHCflurry supported alleles (--list-supported-alleles, Supplementary Table S6) using both MHCnuggets 2.3.2 (74) and MHCflurry 1.4.3 (75) (Supplementary Tables S7 and S8, Supplementary Figures S7-S10). We further cross-referenced these lists of peptides with existing experimentally validated SARS-CoV epitopes present in the Immune Epitope Database (Supplementary Table S4) (76). We then performed consensus binding affinity predictions for the 66 supported alleles shared by all three tools by taking the union set of alleles and filtering for peptide-allele pairs matching the union set of alleles. For the SARS-CoV and SARS-CoV-2 specific distribution of per-allele proteome presentation, we exclude all peptides-allele pairs with >500nM predicted binding. In all cases, we used the netchop v3.0 (77) “C-term” model with a cleavage threshold of 0.1 to further remove any peptides that were not predicted to undergo canonical MHC class I antigen processing via proteasomal cleavage (of the peptide’s C-terminus).

### Global HLA allele and haplotype frequencies

HLA-A, -B, and -C allele and haplotype frequency data were obtained from the Allele Frequency Net Database for 805 distinct populations pertaining to 101 different countries and 2628 distinct major/minor (4-digit) alleles, corresponding to 20,478 distinct haplotypes (https://github.com/pdxgx/covid19). We also identified full HLA genotype data for 3,382 individuals whose HLA types were confined to the 145 HLA alleles studied herein. Population allele and haplotype frequency data were aggregated by country as a mean of all constituent population allele or haplotype frequencies weighted by sample size of the population, but not accounting for representative ethnic demographic size of the population. Global allele frequency maps were generated using the rworldmap v1.3-6 package (78), with total global allele and haplotype frequency estimates calculated as the mean of per-country allele and haplotype frequencies, weighted by each country’s population in 2005.

## Data Availability

Data and code available at https://github.com/pdxgx/covid19. Raw sequence data available through https://www.ncbi.nlm.nih.gov/refseq/.

## ACKNOWLEDGEMENTS

We thank Drs. Christopher Loo and Jeffrey Barnet for their critical reading of the manuscript. We thank Drs. Jonah Sacha and Paul Spellman for their helpful discussions.

## APPENDICES

Appendix 1 contains graphical depictions of sequence alignments for 5 protein sequences (ORF1ab polyprotein, spike, envelope, membrane, and nucleocapsid) across 34 diverse coronavirus proteomes (including all 7 human coronaviruses). Appendix 2 contains a series of world maps displaying the estimated global distributions and frequencies across 145 different HLA alleles, as well as the distribution of predicted SARS-CoV-2 peptide binding and global haplotype frequencies across all haplotypes containing that HLA allele. Appendix 3 contains Supplementary Tables S1-S9S8 as follows: list of SARS-CoV-2 peptides and their predicted binding affinity across 145 HLA alleles (Supplementary Table S1), SARS-CoV-2 peptides conserved across diverse coronavirus sequences (Supplementary Table S2), presence of 8-to 12-mer peptides across four human coronavirus sequences (Supplementary Table S3), prediction of validated SARS-CoV peptides from the Immune Epitope Database (Supplementary Table S4), coronavirus taxonomy and sequence accession numbers for conserved coronavirus proteins (Supplementary Table S4S5), list of 145 HLA alleles used for binding affinity predictions (Supplementary Table S5S6), list of SARS-CoV peptides and their predicted binding affinity across 145 HLA alleles (Supplementary Table S6S7), list of SARS-CoV-2 peptides and their predicted MHCnuggets and MHCflurry binding affinities across 145 HLA alleles (Supplementary Table S7S8), list of SARS-CoV peptides and their predicted MHCnuggets and MHCflurry binding affinities across 145 HLA alleles (Supplementary Table S8S9). Appendix 4 contains the full list of SARS-CoV-2 8-to 12-mers with their individual netMHCpan v4.0 predicted binding affinities across all 145 HLA alleles.

## DISCLAIMER

The contents do not represent the views of the U.S. Department of Veterans Affairs or the United States Government.

## SUPPLEMENTARY DATA

**Supplementary Figure S1:**
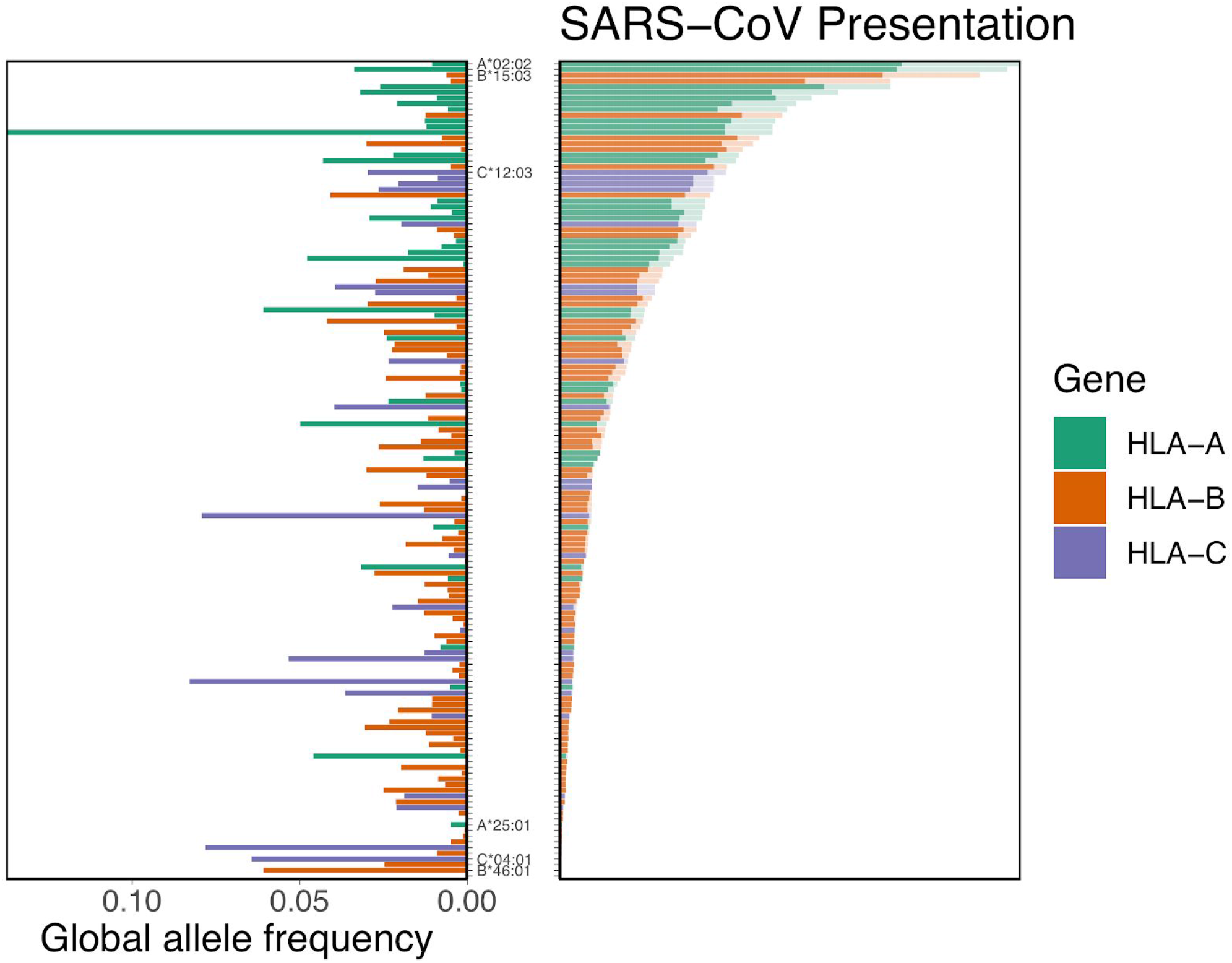
Distribution of HLA allelic presentation of 8-12mers from the SARS-CoV proteome (see Supplementary Table S6). At right, the number of peptides that putatively bind to each of 145 HLA alleles is shown as a series of horizontal bars, with dark and light shading indicating the number of tightly (<50nM) and loosely (<500nM) binding peptides respectively, and with green, orange, and purple colors representing HLA-A, -B, and -C alleles, respectively. Alleles are sorted in descending order based on the number of peptides they bind (<500nM). The corresponding estimated allelic frequency in the global population is also shown (to left), with length of horizontal bar indicating absolute frequency in the population.

**Supplementary Figure S2:**
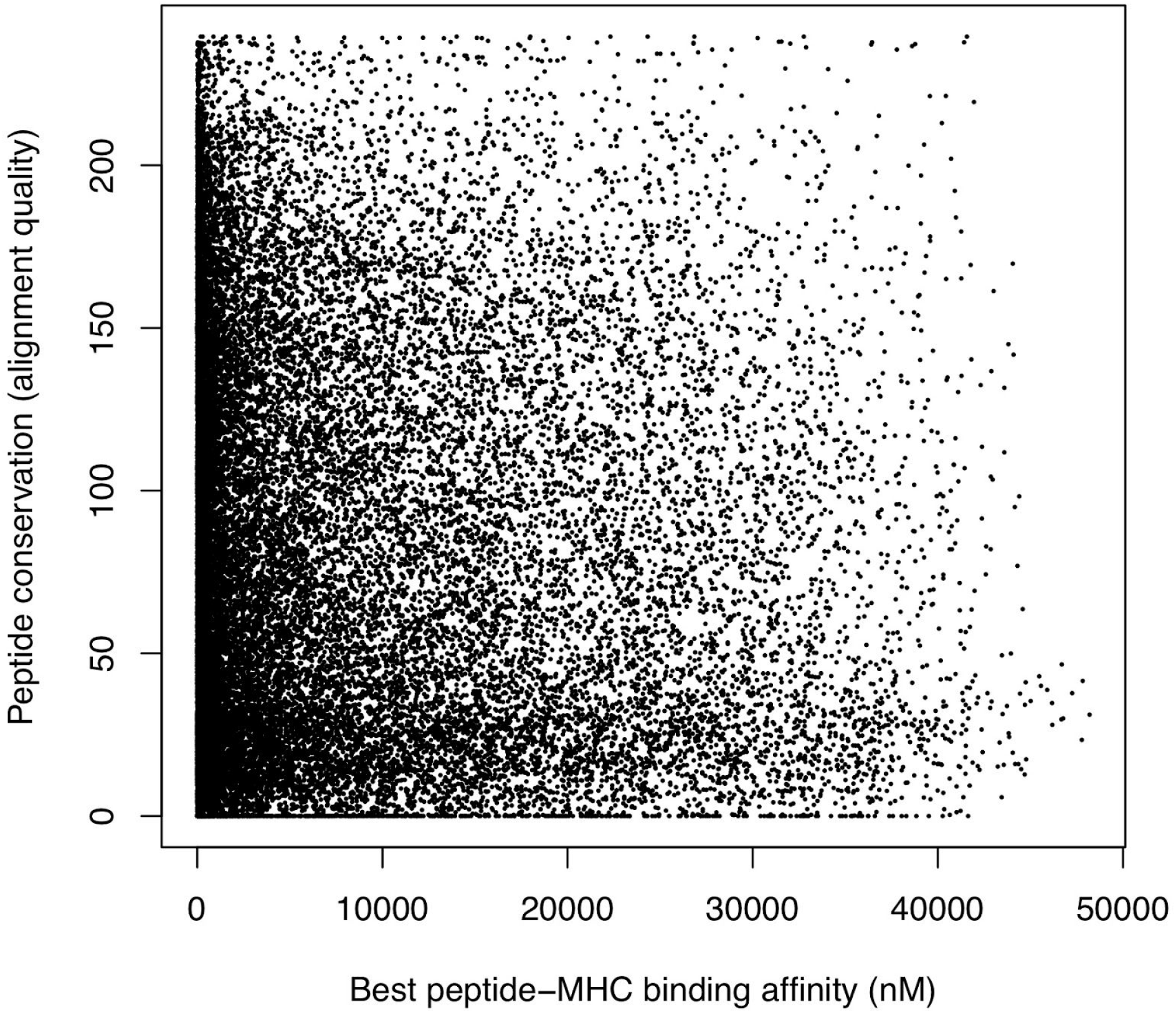
Relationship between predicted peptide-MHC binding affinity and peptide conservation across coronaviruses. Every point represents a single unique peptide covering, together, the entirety of the SARS-CoV-2 proteome. The best predicted MHC binding affinity scores across 145 different HLA alleles are shown for each peptide along the x-axis. Sequence conservation (Clustal Omega alignment score) is shown for each peptide along the y-axis.

**Supplementary Figure S3:**
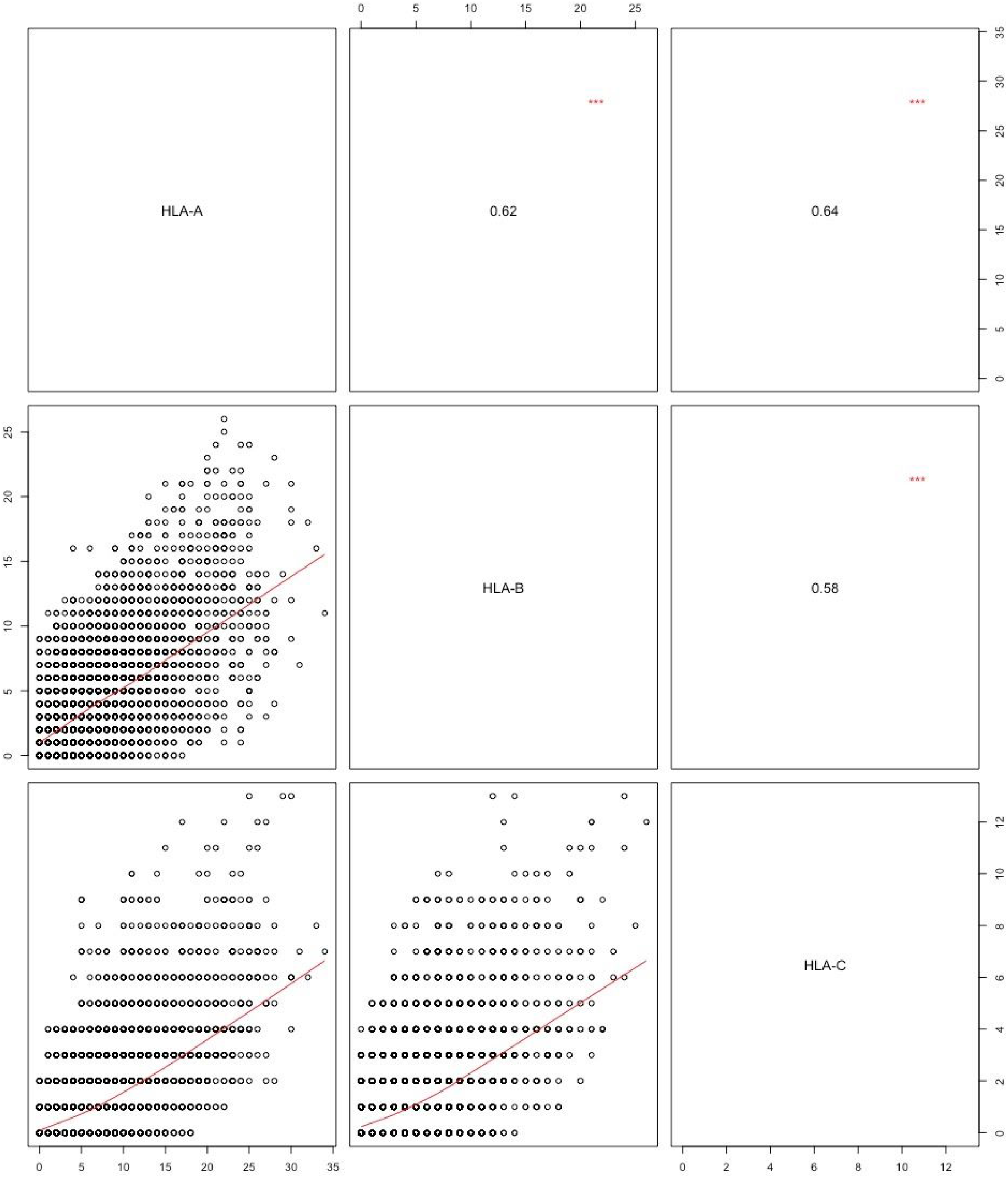
Pairwise relationship of peptide presentation between HLA-A, -B, and -C. In the bottom left three panels, every point represents the pairwise comparison of the number of peptide-allele interactions for all position coordinates. Taken together, the position coordinates cover the entirety of the SARS-CoV-2 proteome. The top right three panels show the quantitative correlation scores between each pair of HLA type comparisons (*** indicates statistical significance).

**Supplementary Figure S4:**
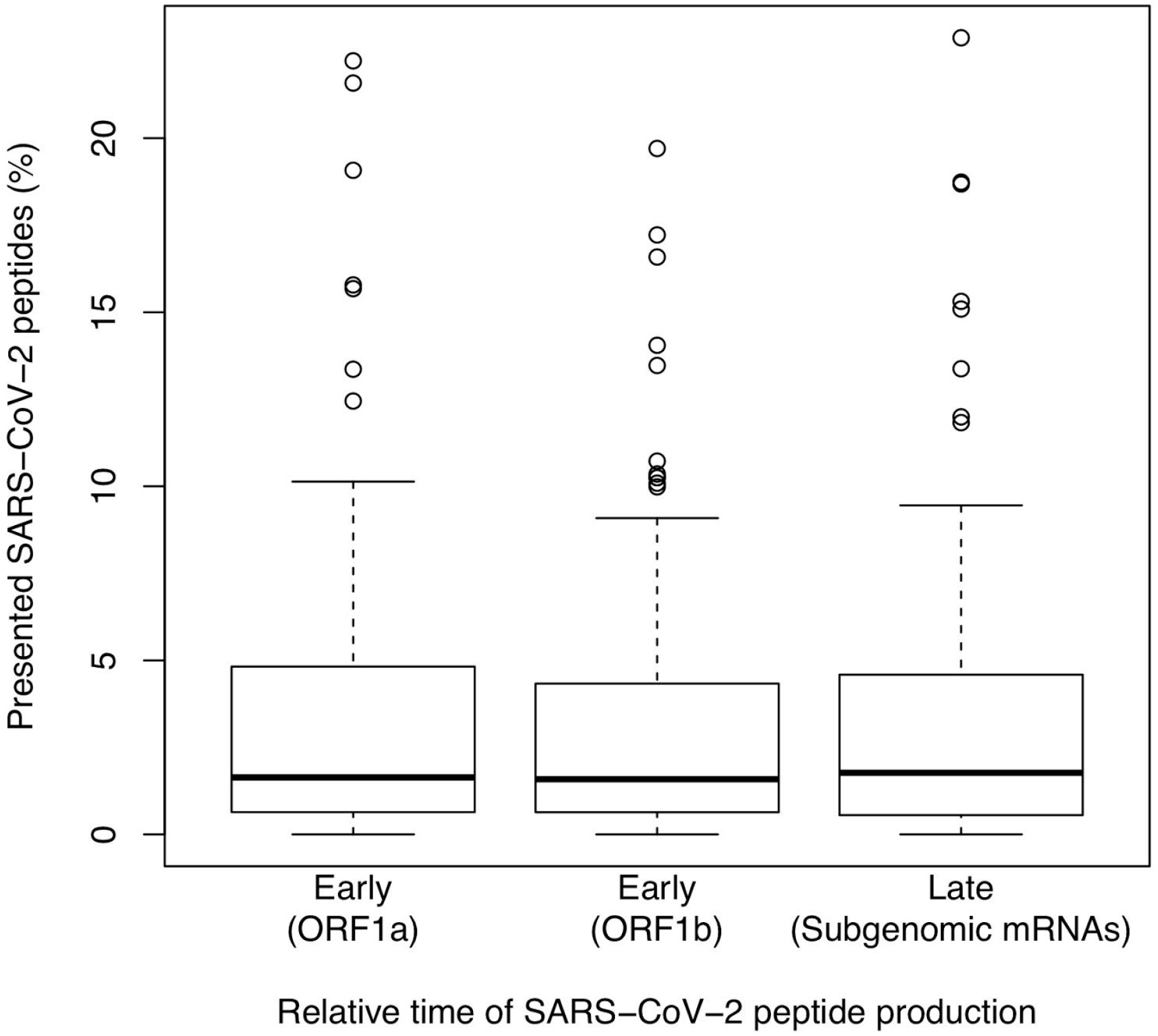
Boxplot distributions of estimated epitope presentation across 145 HLA alleles for early and late SARS-CoV-2 peptides. Capacity for peptide presentation is shown along the y-axis for 145 distinct HLA alleles, for three non-overlapping sets of peptides produced at different timepoints in the viral life cycle as indicated (x-axis). Y-axis percentiles are calculated as the number of peptides from the indicated compartment of the SARS-CoV-2 proteome divided by the total number of presentable 8-to 12-mer peptides from that compartment of the proteome. Dark black lines represent median values, with boxes indicating the 25% and 75% quantiles, with whiskers representing the 25% and 75% quantiles minus or plus the interquartile range, respectively, and with additional outliers shown as open circles.

**Supplementary Figure S5:**
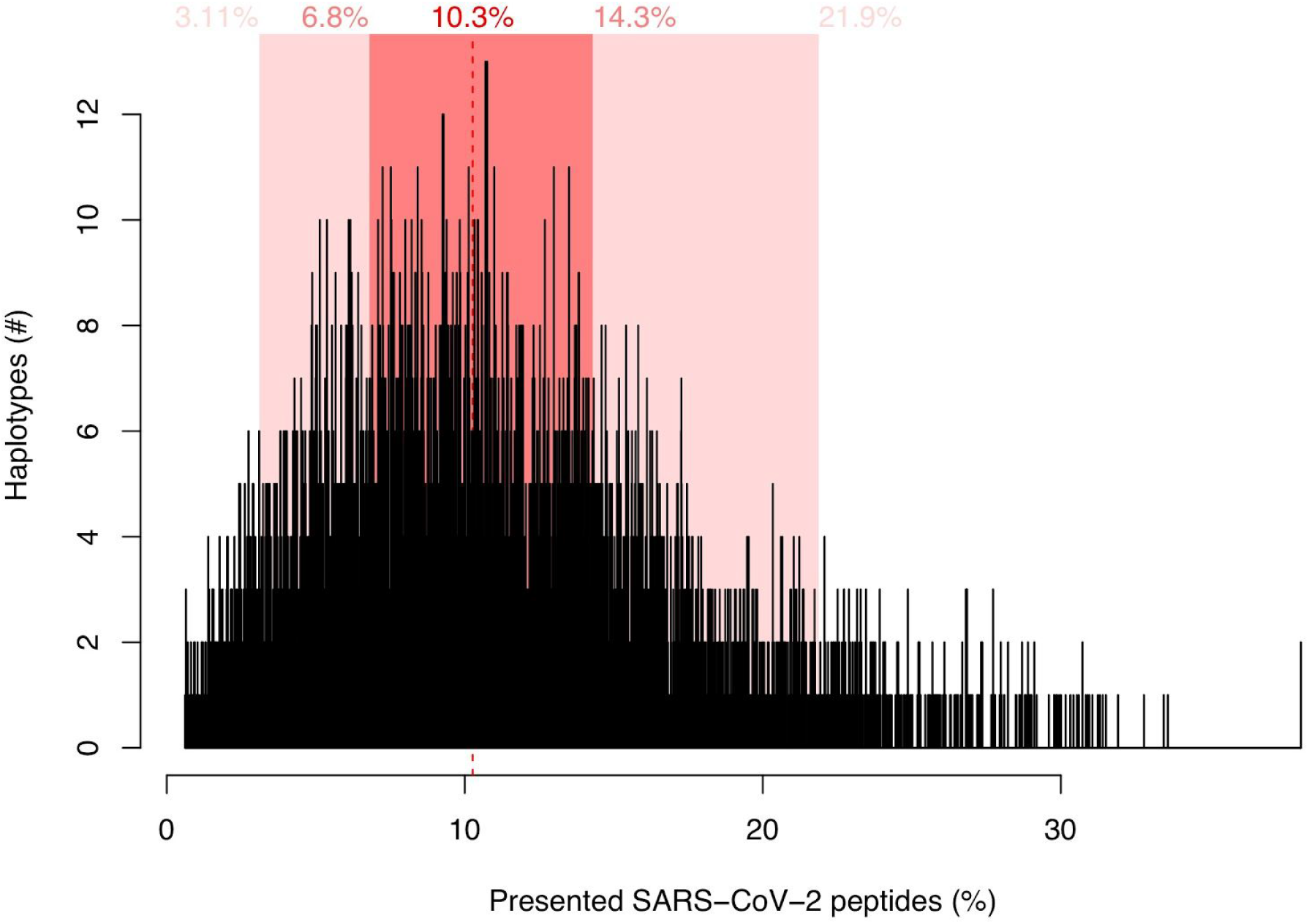
Histogram of SARS-CoV-2 peptide presentation for 5,905 distinct HLA-A/B/C haplotypes. Number of haplotypes are counted along the y-axis, corresponding to their individual capacity (aggregated across all their three component HLA types) to present peptides from the SARS-CoV-2 proteome, shown along the x-axis (percentile is calculated as number of unique peptides presented divided by the total number of presentable 8-to 12-mer peptides from the SARS-CoV-2 proteome). Dashed red line corresponds to the median presentation capacity, while dark and light pink highlighted regions correspond to the 25/75% and 5/95% quantiles, respectively, with numerical values shown in the upper aspect of the plotting region.

**Supplementary Figure S6:**
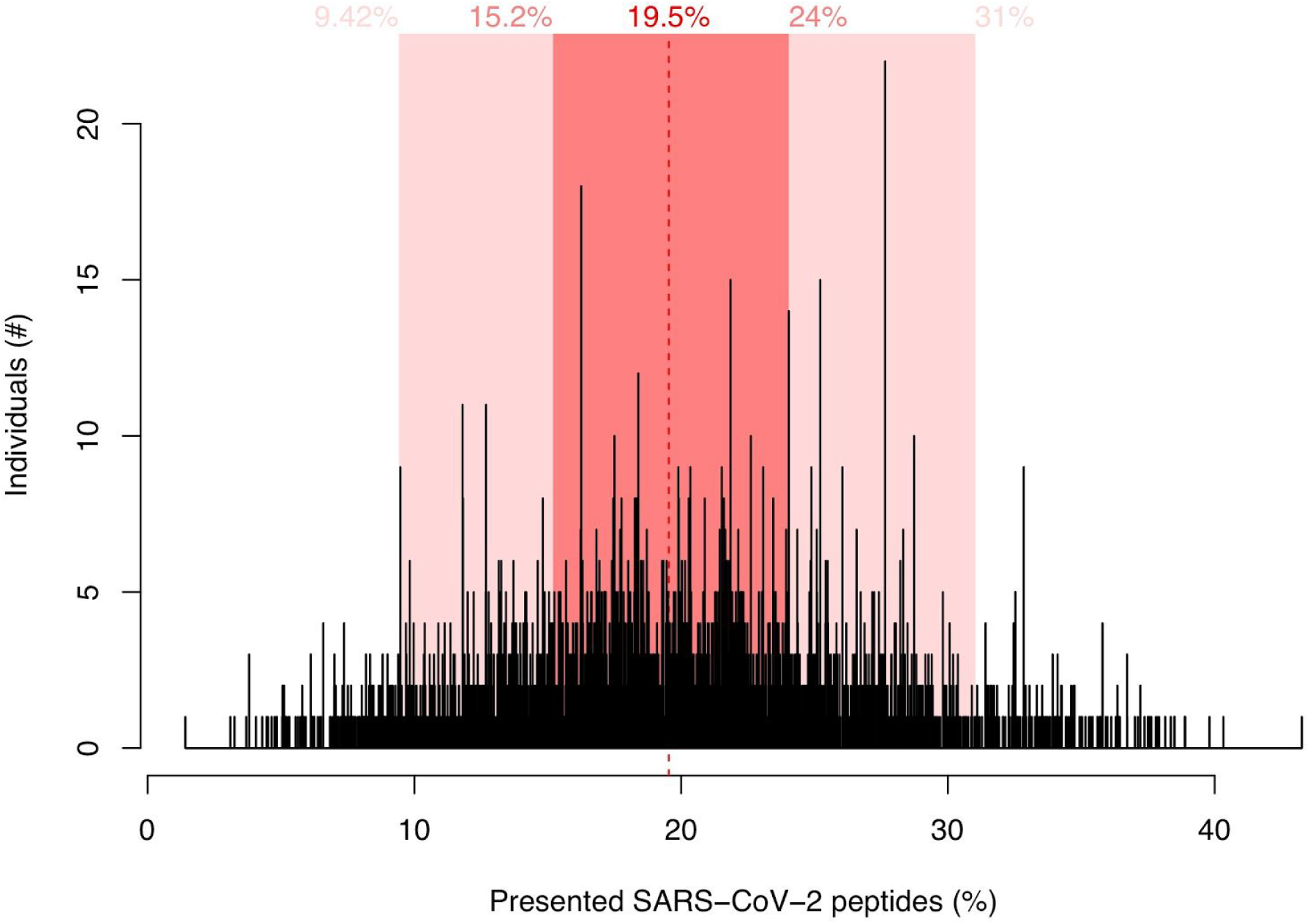
Histogram of SARS-CoV-2 peptide presentation for 3,382 individuals’ full HLA repertoires. Individuals are counted along the y-axis, corresponding to their individual capacity (aggregated across all 6 of their HLA types) to present peptides from the SARS-CoV-2 proteome, shown along the x-axis (percentile is calculated as number of unique peptides presented divided by the total number of presentable 8-to 12-mer peptides from the SARS-CoV-2 proteome). Dashed red line corresponds to the median presentation capacity, while dark and light pink highlighted regions correspond to the 25/75% and 5/95% quantiles, respectively, with numerical values shown in the upper aspect of the plotting region.

**Supplementary Figure S7:**
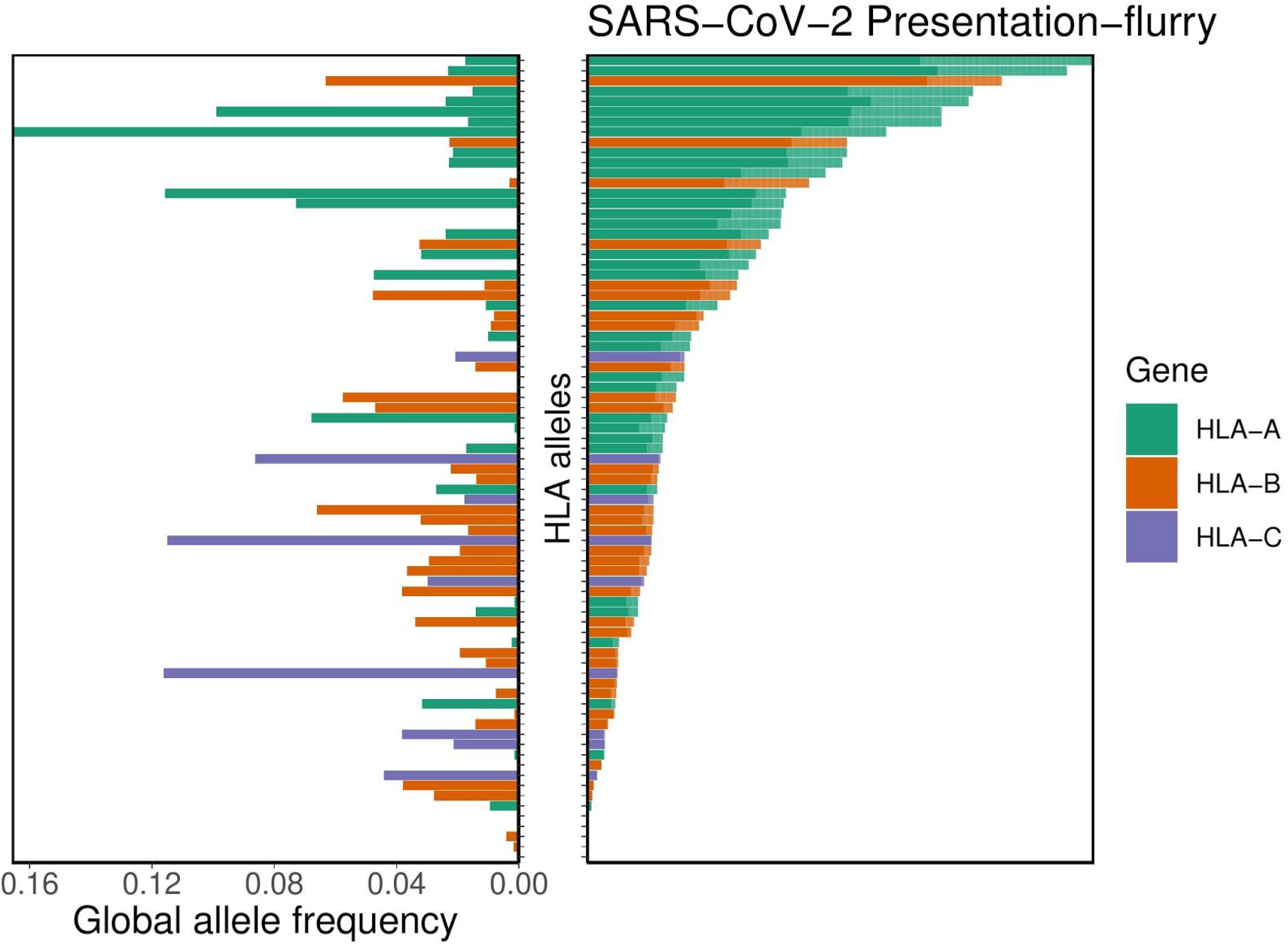
Distribution of HLA allelic presentation of 8-to 12-mers from the SARS-CoV-2 proteome using the tool MHCflurry. At right, the number of peptides (see Supplementary Table S7) that putatively bind to each of 66 HLA alleles is shown as a series of horizontal bars, with dark and light shading indicating the number of tightly (<50nM) and loosely (<500nM) binding peptides respectively, and with green, orange, and purple colors representing HLA-A, -B, and -C alleles, respectively. Alleles are sorted in descending order based on the number of peptides they bind (<500nM). The corresponding estimated allelic frequency in the global population is also shown (to left), with length of horizontal bar indicating absolute frequency in the population.

**Supplementary Figure S8:**
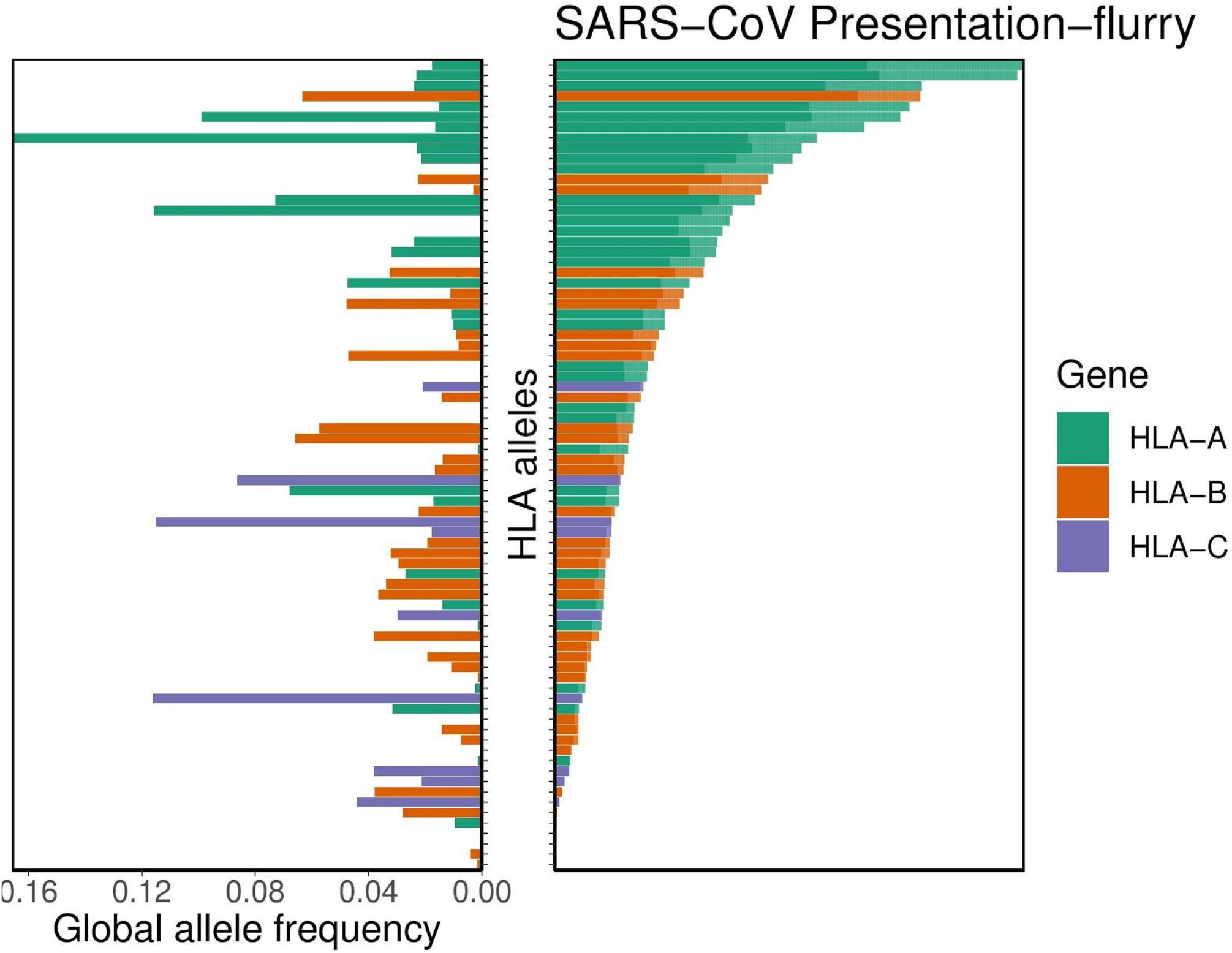
Distribution of HLA allelic presentation of 8-to 12-mers from the SARS-CoV proteome using the tool MHCflurry. At right, the number of peptides (see Supplementary Table S8) that putatively bind to each of 66 HLA alleles is shown as a series of horizontal bars, with dark and light shading indicating the number of tightly (<50nM) and loosely (<500nM) binding peptides respectively, and with green, orange, and purple colors representing HLA-A, -B, and -C alleles, respectively. Alleles are sorted in descending order based on the number of peptides they bind (<500nM). The corresponding estimated allelic frequency in the global population is also shown (to left), with length of horizontal bar indicating absolute frequency in the population.

**Supplementary Figure S9:**
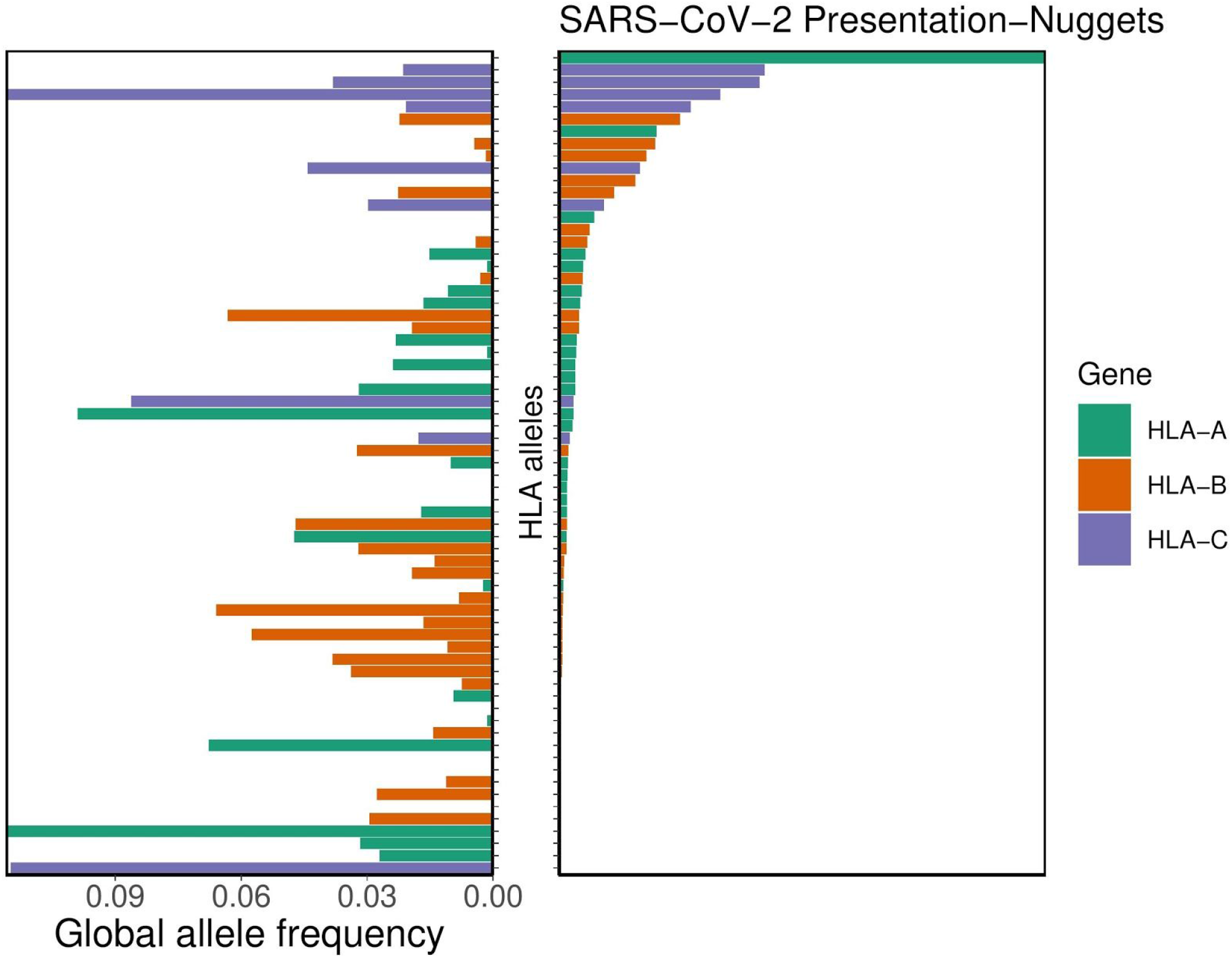
Distribution of HLA allelic presentation of 8-to 12-mers from the SARS-CoV-2 proteome using the tool MHCnuggets. At right, the number of peptides (see Supplementary Table S7) that putatively bind to each of 66 HLA alleles is shown as a series of horizontal bars, with dark and light shading indicating the number of tightly (<50nM) and loosely (<500nM) binding peptides respectively, and with green, orange, and purple colors representing HLA-A, -B, and -C alleles, respectively. Alleles are sorted in descending order based on the number of peptides they bind (<500nM). The corresponding estimated allelic frequency in the global population is also shown (to left), with length of horizontal bar indicating absolute frequency in the population.

**Supplementary Figure S10:**
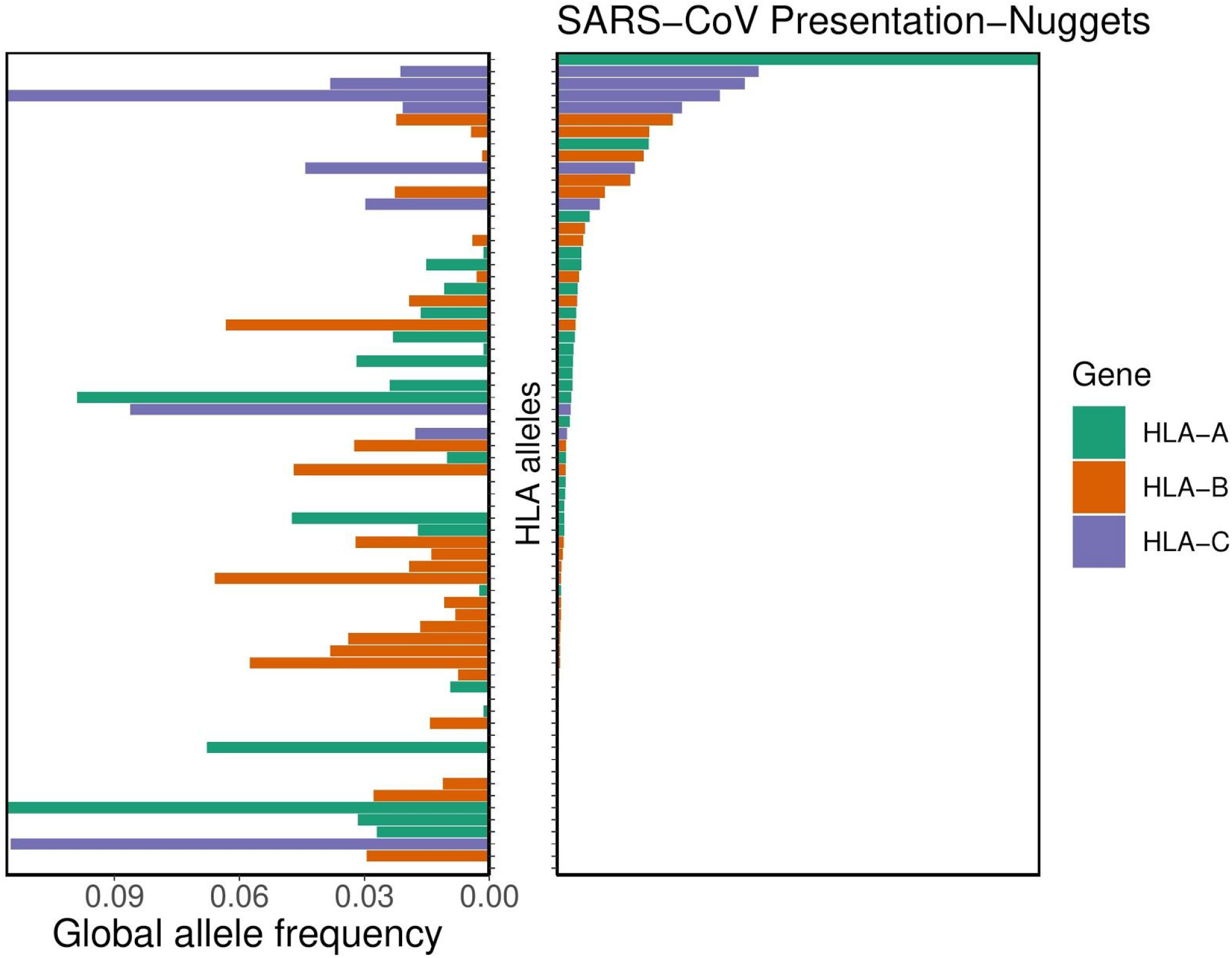
Distribution of HLA allelic presentation of 8-to 12-mers from the SARS-CoV proteome using the tool MHCnuggets. At right, the number of peptides (see Supplementary Table S8) that putatively bind to each of 66 HLA alleles is shown as a series of horizontal bars, with dark and light shading indicating the number of tightly (<50nM) and loosely (<500nM) binding peptides respectively, and with green, orange, and purple colors representing HLA-A, -B, and -C alleles, respectively. Alleles are sorted in descending order based on the number of peptides they bind (<500nM). The corresponding estimated allelic frequency in the global population is also shown (to left), with length of horizontal bar indicating absolute frequency in the population.

